# How much disease-risk is due to old age and established risk factors?

**DOI:** 10.1101/2023.01.20.23284838

**Authors:** Anthony J. Webster

**Affiliations:** Nuffield Department of Population Health, Lindgren Group, Big Data Institute, Old Road Campus, University of Oxford, Oxford, OX3 7LF, UK

## Abstract

As improved healthcare leads to older populations, individuals will increasingly experience multiple diseases, possibly concurrently (multimorbidity). This article explores whether age and established risk factors are sufficient to explain the incidence rates of multiple, possibly coexisting diseases. By accounting for the limited age-range in UK Biobank data, previous work demonstrated that a Weibull model could accurately describe the incidence of ∼60% of the most common primary hospital diagnoses of diseases. These are used here to predict the age-dependent incidence of diseases with adjustment for established risk factors. A “Poisson binomial” model is combined with these to predict the total number of occurrences of each disease in the UK Biobank cohort that would be expected without pre-existing (prior) disease. For 123 diseases in men and 99 diseases in women, the total observed new cases of each disease (including those from individuals with pre-existing diseases and multimorbidity), were found to be approximately 1.5 times greater than that predicted for individuals without prior disease, and could not be explained by natural statistical variation. The multiple of 1.5 was sufficiently consistent across different diseases to prevent its use for classification of disease types, but there were differences for sub-groups such as smokers with high body mass index, and for some classes of disease (as defined by the International Classification of Diseases version 10). The results suggest that empirical modelling might allow reliable predictions of primary causes of hospital admissions, helping to facilitate the planning of future healthcare needs.

---

Multi-morbidity is increasingly common in developed economies such as the United Kingdom. It is widely believed to involve clusters of diseases and that disease risks are modified by underlying conditions [1–4]. Despite considerable work to characterise multimorbidity in terms of clusters of diseases (e.g. see the recent review [4]), there have been comparatively few recent studies to determine the influence of pre-existing conditions on the age-dependent incidence rates of disease [2, 5–11]. Here we restrict attention to primary hospital diagnoses of common diseases, and consider whether the number of disease types that occur in individuals are consistent with expectations based on age and established risk factors. We use results from a recent study [12] that used a Weibull distribution to explore whether the age-related incidence of diseases in UK Biobank [13] are consistent with multistage disease processes [12, 14–16]. Of the 800 common diseases that were considered, 450 were consistent with the model, and there were sufficient cases for 172 (or 156) to be modelled with adjustment for 7 (or 9) established risk factors in men (or women) [12]. By combining these results with a “Poisson Binomial” distribution [17, 18], this article assesses whether differences in disease incidences are different to that expected from natural statistical variation. An outline of the analysis is in figure 1. The statistical method is described next, the results are presented and discussed in the following two sections, and the conclusions summarise the main results.

**Figure 1:**
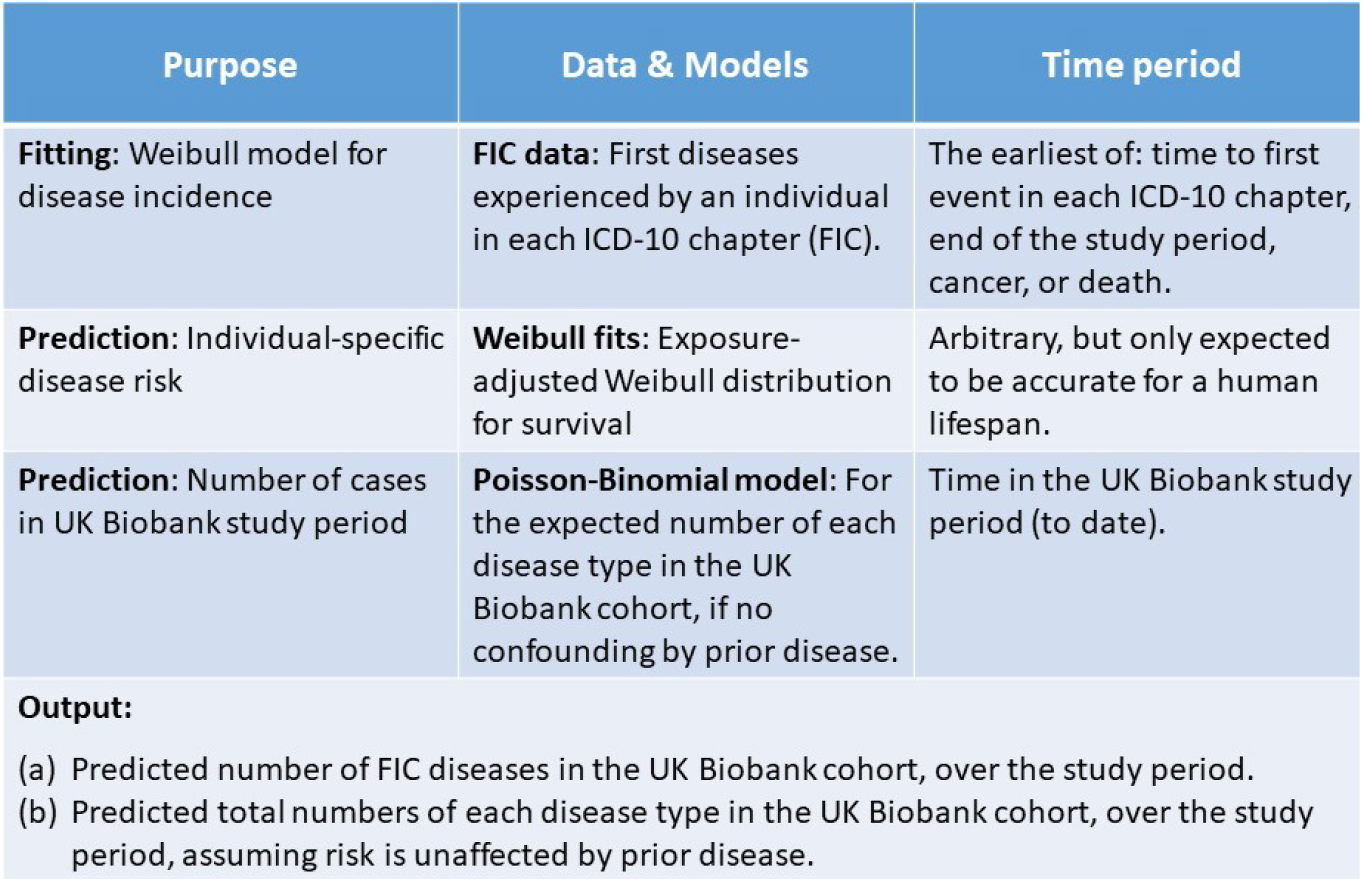
The table summarises the data and analysis methods used (top to bottom).

## Independent disease incidence rates

### Statistical background

We test the null hypothesis that the incidence rate of each disease type, is independent of the presence of different previous, or co-existing diseases. Using incidence rates of each disease type, that are estimated for a scenario that approximates no prior disease, we calculate the probability of observing *N*_*j*_ cases of disease type *j* in the UK Biobank cohort when there is no prior disease. Let *S*_*ij*_(*t*) be the probability of person *i* surviving disease *j* until age *t*, then 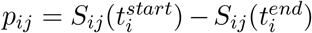 is the probability of person *i* first experiencing disease *j* between the ages 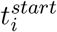 and 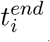 during which they were in the study. Let *X*_*ij*_ = 1 if individual *i* has disease *j*, and zero otherwise. Then if disease risks are independent, the probability of *i* = 1 to *i* = *n* individuals observing the set of diseases {*X*_*ij*_}, is,

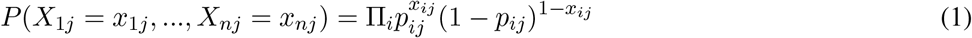

The probability of observing *N*_*j*_ = Σ_*i*_ *X*_*ij*_, where *N*_*j*_ is the number of individuals who experience disease of type *j*, is given by the “Poisson Binomial” distribution,

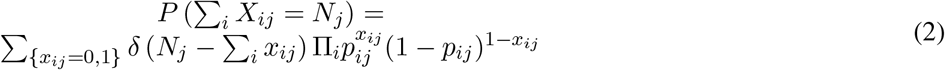

where *δ*(*s*) is the Dirac delta function that equals 1 when *s* = 0 but is zero otherwise, and the sum is over all values *x*_*ij*_ = 0 and 1, for *i* = 1..*n*. The mean and variance of the distribution are,

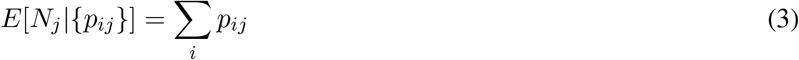

and,

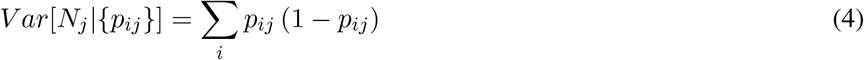

Simple derivations of these results are given in the Materials and Methods. For *p*_*ij*_ ≪1, as is the case for most of the diseases in most individuals considered here [12, 19, 20], then the distribution will approximate a Poisson distribution, which provides another reason why the Poisson distribution is common - it approximates the distribution for the number of events arising from rare independent processes.

For each disease *j* and individual *i*, the multivariate *δ*-method [21] allows variances *σ*_*ij*_ to be estimated for the probability 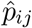 of disease *j* during the time observed, with 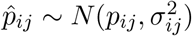. The law of total variance states that,

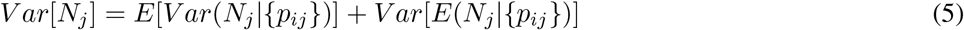

that can be evaluated using Eqs. 3 and 4. Surprisingly perhaps, the variances 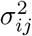 that arise from integrals involving 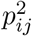 cancel, and do not appear in the result, that is,

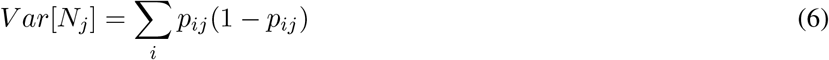

and can be estimated by “plugging in” the maximum likelihood estimates 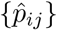 for {*p*_*ij*_}. In practice the estimated variances 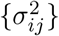 were important for “quality control”, allowing the identification of poor quality estimates that are too imprecise, or unlikely to satisfy the assumptions needed for application of the *δ*-method. Specifically, diseases were excluded if estimates for parameters *x* = *k* or *x* = *L* had *s*.*e*.(*x*)*/x >* 0.5, or where the *δ*-method failed to give a numeric estimate, and it was confirmed that the remaining estimates had 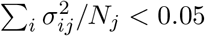. The negative correlation between parameters *k* and *L* led to smaller variances for estimates than might have been expected based on estimates for the variances of *k* and *L* individually.

### Data analysis

Diseases included in the study were defined as a collection of one or more 3- and 4-digit disease codes from the International Classification of Diseases Version 10 (ICD-10) [22], that were selected by three epidemiologists with individual backgrounds in pathology, general practice, and statistics, based on a set of predetermined criteria as detailed previously [23]. Diagnoses used primary cause of admission in hospital episode statistics, that ensured that the diseases had passed a threshold of severity prior to diagnosis. The study period for an individual was between joining the study and 31st January 2020, after which admission rates will be influenced by the Covid pandemic. To minimise the potential influence of prior diseases or medications, individuals were excluded if they had a prior diagnosis of a cancer other than non-melanoma skin cancer before the study started, or a report of cardiovascular disease (either self-reported or in hospital records).

The study considered all primary diagnoses within the study period (“all cases”), and similar to previous work [12, 19, 23], the first primary diagnosis that an individual receives in each ICD-10 chapter (“FIC”). By considering only the first diagnosis in each ICD-10 chapter, the intention was to minimise confounding by prior disease, while maximising the number of cases that are included in the study. Importantly, because the study considers the number of different diseases experienced by an individual, counts do not include repeat admissions for the same disease in the same individual.

A previous study of UK Biobank data identified 450 diseases whose FIC incidence rates could be modelled by a Weibull distribution [12],

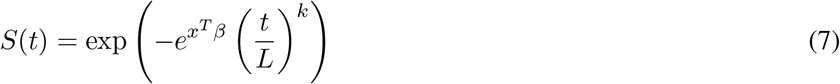

where *t* is age, *k* and *L* are parameters, and *x, β* are vectors of covariates and parameters. Individuals were excluded if they had a report of the disease before the study’s start. Maximum likelihood estimates and their covariances were calculated by left-truncating at the age when participants joined the study, taking age of event as the age of disease-onset, and right-censoring if there is cancer, death or the study ends before disease onset. Adjustment was for the established risk factors of smoking (never, previous, current), diabetes (yes, no), alcohol (rarely - less than 3 times per month, sometimes - more than 3 times per month but less than 3 times per week, regularly - more than 3 times per week), deprivation tertile (min, mid, max), education (degree level, post-16, to age 16), and sex-specific tertiles of height and BMI (min, mid, max). Baseline was never smoked, no diabetes, sometimes drink, min deprivation, degree-level education, middle BMI, minimum height and deprivation tertiles, and women with no HRT use or children.

For each disease whose age-dependent incidence could be modelled sufficiently well, the observed number of cases in the UK Biobank cohort was plotted versus the number of predicted cases assuming no confounding by prior disease (Figure 2). The results for men and women were plotted separately, and excluded diseases whose predicted case numbers differed by more than 4 standard deviations (sd) from the observed numbers of first diseases in each ICD-10 chapter (with minimal confounding by prior disease). The (lenient) threshold of 4 sd was intended to include as many diseases as possible, while limiting the risk of outliers from substantially influencing subsequent results. In practice, for most of the diseases, 4 sd was found to be much less than the differences between the number of observed and predicted cases.

**Figure 2:**
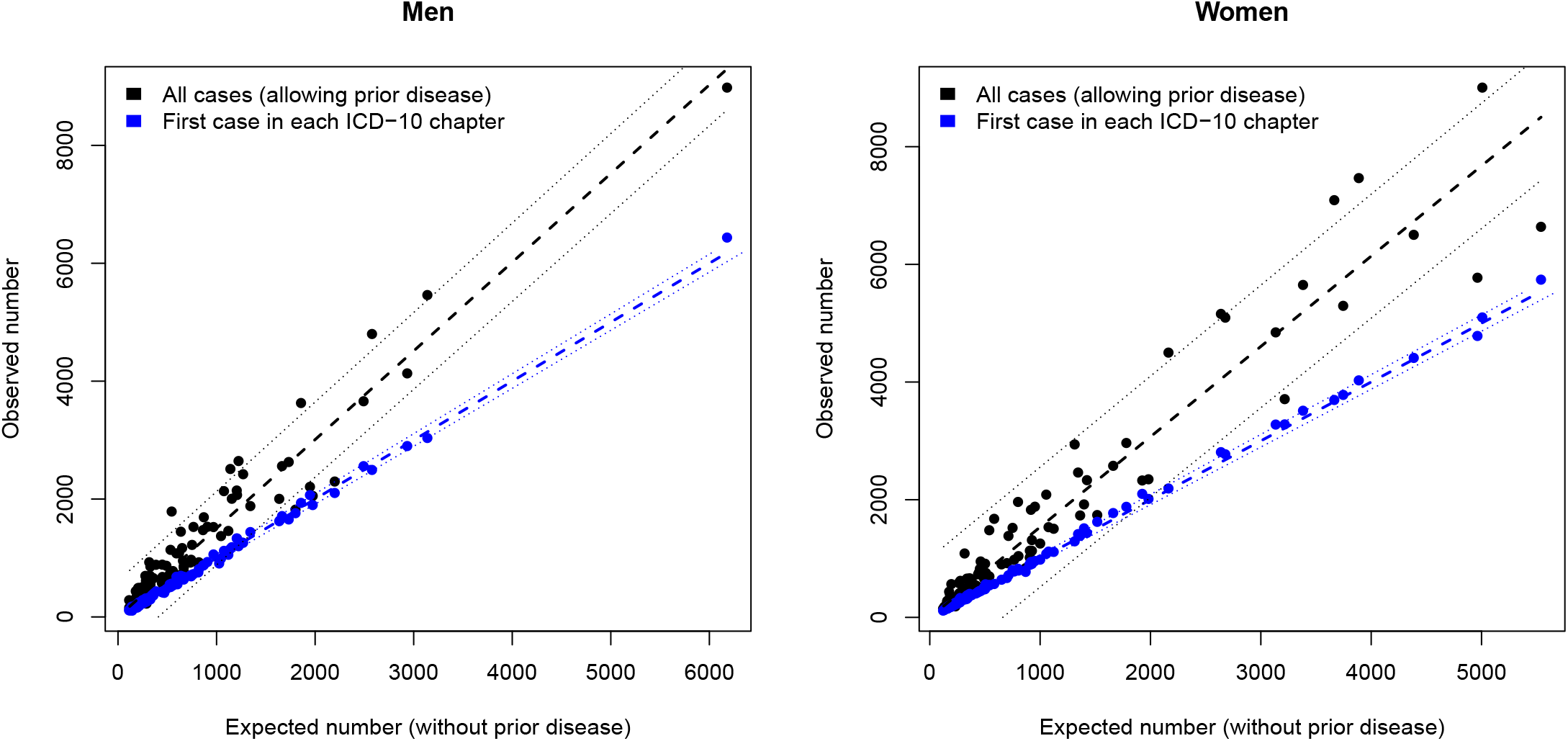
For each disease, the number of individual incidences are predicted in the absence of prior disease (Expected number), and plotted against the total observed number of cases (black) and observed number of cases where the disease is the first an individual experiences in each ICD-10 chapter (blue). 95% confidence intervals are estimated using Eq. 6 as *±*1.96 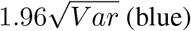, and for predictive confidence intervals of the linear fit (black).

Confidence intervals are reported as *±*1.96*s*, where *σ* is the estimated standard deviations of the maximum likelihood estimate (MLE). For an MLE 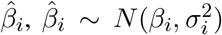, and tests for equality of MLEs use 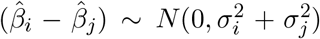, so statistically significant differences at the 0.05 level will have 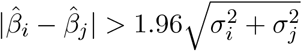.

## Results

Figure 2 plots the number of each disease type observed in the UK Biobank cohort, versus the predicted number of cases if there is no confounding by prior disease, with poorly modelled data removed (as described above). A straight-line fit through the origin estimated the number of observed diseases in men to be 1.50 [1.45,1.55] times the expected number of diseases without prior disease, and 1.53 [1.47,1.59] in women (tables 1 and 2). There is no statistically significant difference between the fits for men and women. The R-squared value for both fits was 0.96. Data for individual diseases are given in the Supplementary Material.

**Table 1:**
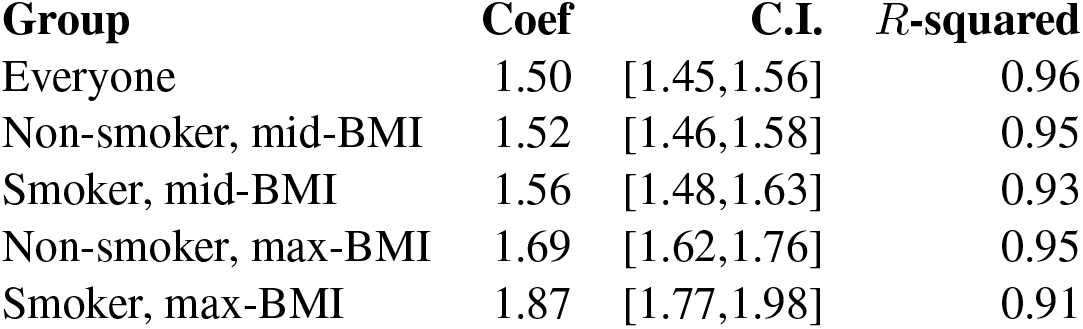
Increased incidence associated with prior disease in men. The estimated slope (Coef), its confidence intervals (C.I.), and *R*-squared coefficients for Everyone (Figure 2), and sub-groups.

**Table 2:** Increased incidence associated with prior disease in women. The estimated slope (Coef), its confidence intervals (C.I.), and *R*-squared coefficients for Everyone (Figure 2), and sub-groups.

| Group | Coef | C.I. | *R*-squared |
| --- | --- | --- | --- |
| Everyone | 1.53 | [1.47,1.6] | 0.96 |
| Non-smoker, mid-BMI | 1.42 | [1.36,1.48] | 0.96 |
| Smoker, mid-BMI | 1.60 | [1.51,1.69] | 0.92 |
| Non-smoker, max-BMI | 1.61 | [1.52,1.70] | 0.94 |
| Smoker, max-BMI | 1.74 | [1.61,1.87] | 0.88 |

Subgroups that were expected to include the highest and lowest risk groups were also considered (tables 1 and 2), these included: non-smokers in the mid-BMI tertile, smokers in the mid-BMI tertile, non-smokers in the max-BMI tertile, and smokers in the max-BMI tertile. Statistically significant differences between estimates for men and women (at the 0.05 level), are only found for the non-smoking, mid-BMI group, that have lower estimates for females. For both men and women there were statistically significant differences between the group including everyone, and the subgroup of smokers in the max-BMI tertile. For men only, there were statistically significant differences between the group including everyone, and non-smokers in the max-BMI tertile. Overall the increase in disease rates above those expected without prior disease, tended to be higher for groups that would already be expected to have higher disease risk (e.g. smokers and, or, the top BMI tertile).

The ICD-10 coding system groups disease hierarchically into chapters that are intended to capture the dominant disease types such as cancers (chapter III) or cardiovascular diseases (chapter IX) for example. There were comparatively few diseases representing some ICD-10 chapters (see figure 3), but with that caveat, figure 3 indicates which disease risks appear to be most susceptible to prior disease. Some chapters such as diseases of the ear (VIII), or of the nervous system (VI), had case numbers similar to that expected without prior disease. In contrast, diseases of the digestive system (XI), circulatory diseases in men (IX), genitourinary diseases (XIV), musculoskeletal diseases (XIII), and unclassified symptoms of potentially unknown origin (XVIII), have case numbers that are all increased in risk by a factor of more than 1.5 above that expected in someone without pre-existing disease.

**Figure 3:**
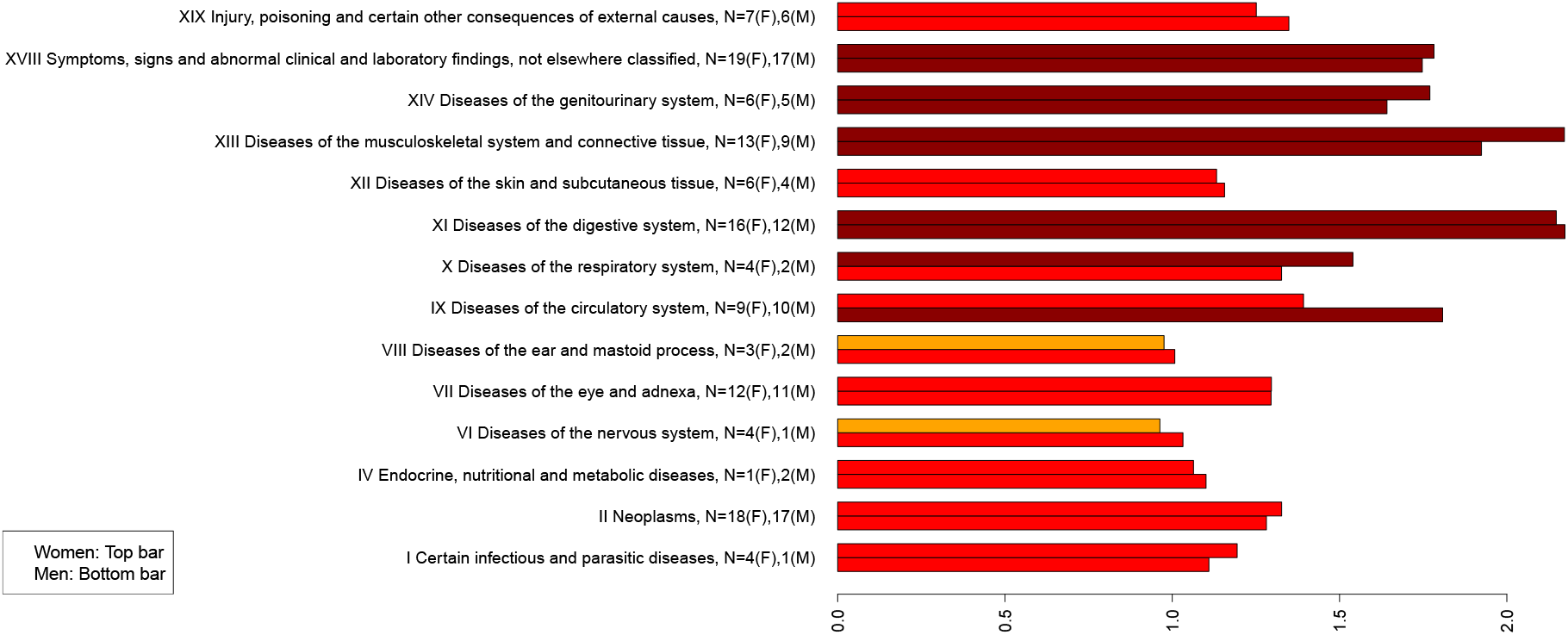
The mean increase in observed cases of disease above that predicted without prior disease, plotted by ICD-10 chapters (women top, men bottom). Numbers of diseases in each bar are indicated by N, with (F) for women and (M) for men. Data for individual diseases are in the Supplementary Material.

## Discussion

It had originally been expected that prior disease would increase disease rates, and that these increases would vary substantially between disease types. This could have allowed diseases to be characterised and classified by their sensitivity to prior disease. Instead the increases in disease risk were similar and evenly spread across the diseases considered, and between both men and women. This raises the possibility of using empirical modelling to link estimated disease rates without prior disease that are estimated by conventional epidemiological studies, to those that are observed in practice. Furthermore, in contrast to methods that use present trends to predict future demand [6], the (risk-factor adjusted) models here can be used in counterfactual modelling [24–26] of potential interventions.

There were however, statistically significant differences between the increased risks in subgroups of individuals such as smokers with high BMI, and the cause of these differences are unclear. It could be that the risk factors are increasing both the underlying disease risk (without prior disease), and the influence of prior disease on your subsequent disease risk. Alternately, the increase in underlying disease risk could lead to a greater number of co-existing disease conditions, that together increase disease risk more than for someone with fewer prior diseases. The particular diseases and mechanisms by which prior disease modifies future disease risk need to be identified and understood. Nonetheless, figure 2 suggests that your overall disease risk is driven by your underlying (disease-free) disease risk, and reducing well-understood disease risks will reduce your overall expected burden of disease in old age.

Figure 3 clearly suggests differences in susceptibility of disease risk to prior diseases, based on existing ICD-10 classifications. However, no obvious clusters of diseases were found based on the increased rate of disease, although it is possible that differences could develop if more diseases were able to be included in the study. The figure also suggests that predictions for observed disease rates might be improved by considering predictions within ICD-10 chapters.

## Limitations

This study considered data in UK Biobank, and approximately 60% of the 400 most common diseases whose incidence rates could be modelled accurately with a Weibull distribution [12]. Of these, more stringent fitting requirements detailed in the Methods reduced the number of diseases considered to 123 in men and 99 in women (222 in total). Future work may be able to improve the modelling of disease onset rates, that would allow a more comprehensive study. In addition, the UK Biobank cohort is known to be a poor representation of the UK population as a whole, and it is possible that results might differ for a different cohort. For these reasons, the estimates in figure 2 and tables 1, 2, are likely to be modified for analyses with different cohorts and different collections of diseases. Studies in different cohorts, and with different periods of follow-up, will be needed to explore the generality of these results.

The main purpose of this article was to explore how expected disease rates due to established risk factors and old age, are modified by prior disease. Future work will be needed to establish the diseases and disease-mechanisms that are responsible for modifying subsequent disease risk. Drug use and poly-pharmacy are also likely to be important. It is certain that some medications, such as statins, can modify risks of diseases other than those that they are primarily intended to treat.

## Conclusions

By combining a Poisson-Binomial distribution with a Weibull model for the age-related incidence of disease, this study considered how much disease risk in the UK Biobank cohort could be explained by age and established risk factors. For the 222 diseases considered, it was found that disease rates were much greater than would be expected without prior disease, or than could be explained by natural statistical variation. The increases in risk were comparatively uniform across the diseases considered, and between men and women, but appeared to differ between ICD-10 classifications of disease types and for subgroups such as smokers with high BMI. However, there were no obvious clusters of diseases in terms of increases in disease risk, that was found to be approximately 1.5 times those expected of individuals without prior disease. In other words, for the 222 diseases considered, approximately two thirds of primary hospital admissions in the UK Biobank cohort were as expected based on age and established risk factors, with the additional third presumed to involve an increased risk that is directly or indirectly associated with prior diseases.

More generally, if the overall disease risks were (approximately) proportional to disease risks without prior disease, there would be several important implications. Firstly, avoiding known risk factors for disease would also reduce the risk, or delay the onset, of multiple diseases in old age. Secondly, it would reaffirm the value of conventional epidemiological studies of disease risk that deliberately avoid potential confounding by prior disease. Thirdly, because the (risk-factor adjusted) model can extrapolate beyond the end of the study period, and the confidence intervals in figure 2 are all comparatively narrow, reliable empirical modelling of future disease rates might be possible. This would be helpful for future healthcare planning.

Further studies are needed to increase the breadth of diseases that can be modelled, to explore the generality of the results in other cohorts, and to test the model’s predictive performance. Despite the present limitations, the results provide the first quantitative characterisation of how prior disease modifies the incidence rates of a wide range of disease types, and a methodology that can be used or developed in future studies.

### The Poisson-Binomial model

Simple derivations of the key properties of the “Poisson Binomial” distribution are outlined below, with comprehensive proofs given elsewhere [17, 18].

As discussed in the main text, the probability of observing *N*_*j*_ = ∑_*i*_ *N*_*ij*_, where *N*_*ij*_ is the number of different diseases observed in individual *i*, is given by the “Poisson Binomial” distribution,

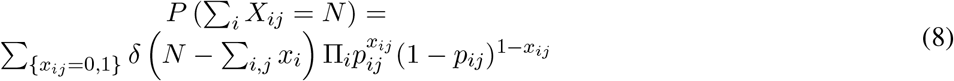

where *δ*(*s*) is the Dirac delta function that equals 1 when *s* = 0 but is zero otherwise, and the sum is over all *x*_*ij*_ for *i* = 1..*n*.

The generating function for 8 is,

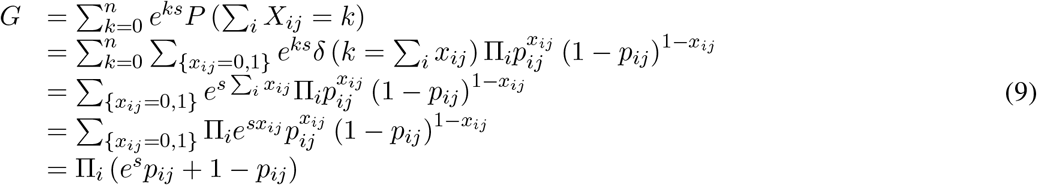

where Eq. 8 was included in the second line, the Dirac delta-function led to *e*^*ks*^ being replaced by *e*^*k*^Σ_*i*_ *x*_*ij*_ when summing over *k* in the third line, the fourth line includes *e*^*k*^Σ_*i*_ *x*_*ij*_ within the product over *i* and *j*, and the final line has summed over each *x*_*ij*_ taking values of 0 and 1 with the sum’s evaluation seen most easily by explicitly considering *i* = 1 and factorising the result. Moments of Eq. 8 can be obtained by taking derivatives of Eq. 9 with respect to *s*, and then evaluating them at *s* = 0, with for example,

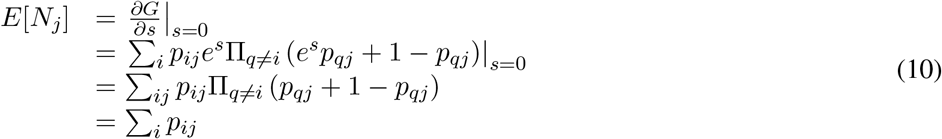

Similarly the second moment is,

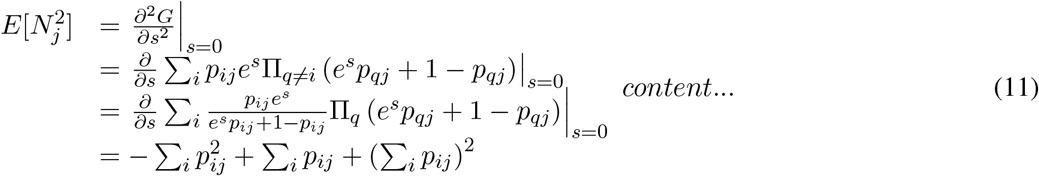

where in the final line the first term arises from the derivative of the denominator of the first term in line 3, the second term from the derivative of the numerator of the first term in line 3, and the final term is from the derivative of the product in line 3 similarly to evaluating *E*[*N*_*j*_]. Combining Eq. 10 with Eq. 11, the variance can be evaluated as,

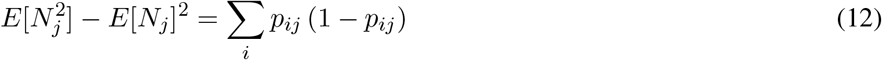

*E*[*N*_*j*_] and *V ar*[*N*_*j*_] can alternatively be written as 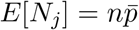 and 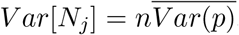, where the bars denote averages. If *p*_*ij*_ ≪ 1, then the variance Eq. 12 has *V ar*[*N*_*j*_] ≃ ∑_*i*_ *p*_*ij*_ = *E*[*N*_*j*_], as would be the case for a Poisson distribution with rate *λ*_*ij*_ = ∑_*i*_ *p*_*ij*_ and 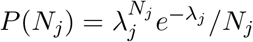. In fact, when *p*_*ij*_ ≪ 1, the Poisson-Binomial distribution tends to the Poisson distribution [17], as can be seen from the moment generation function Eq. 9, that approximates the Possion distribution for *p*_*ij*_ → 0. Using Eq. 9 and expanding in terms of *p*_*ij*_,

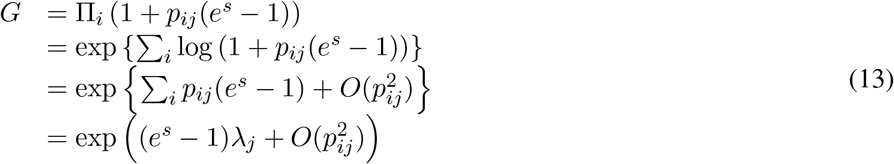

where *λ*_*j*_ = ∑_*i*_ *p*_*ij*_, and the generating function for a Poisson distribution is exp ((*e*^*s*^ − 1)*λ*_*j*_). The above argument suggests that provided *p*_*ij*_ ≪1, then a Poisson distribution can model the number of events due to *n* independent processes that each have different but low probabilities *p*_*ij*_ of occurring.

## Data Availability

UK Biobank data can be accessed by application through www.ukbiobank.ac.uk.

## Code and data availability

UK Biobank data can be accessed by application through www.ukbiobank.ac.uk. UK Biobank has approval by the Research Ethics Committee (REC) under approval number 16/NW/0274. UK Biobank obtained participant’s consent for the data to be used for health-related research, and all methods were performed in accordance with the relevant guidelines and regulations. R code used to produce figures from summary data will be made available from https://osf.io. R packages used in this study include survival[27], grr[28], data.table[29], and maxLik[30].

## Acknowledgments

This research has been conducted using data from UK Biobank, a major biomedical database, under application number 42583. This research was supported by an intermediate research fellowship from the Nuffield Department of Population Health (NDPH), University of Oxford.

## References

[1] Karen Barnett, Stewart W. Mercer, Michael Norbury, Graham Watt, Sally Wyke, and Bruce Guthrie. Epidemiology of multimorbidity and implications for health care, research, and medical education: a cross-sectional study. Lancet, 380(9836):37–43, JUL 7 2012.

[2] Bruno Pereira Nunes, Thayna Ramos Flores, Gregore Iven Mielke, Elaine Thume, and Luiz Augusto Facchini. Multimorbidity and mortality in older adults: A systematic review and meta-analysis. Archives of Gerontology and Geriatrics, 67:130–138, NOV-DEC 2016.

[3] C.J.M. Whitty and F.M. Watt. Map clusters of disease to tackle multimorbidity. Nature, 579:494–496, 2020.

[4] Soren T. Skou, Frances S. Mair, Martin Fortin, Bruce Guthrie, Bruno P. Nunes, J. Jaime Miranda, Cynthia M. Boyd, Sanghamitra Pati, Sally Mtenga, and Susan M. Smith. Multimorbidity. Nature Reviews Disease Primers, 8(1), JUL 14 2022.

[5] Concepcio Violan, Quinti Foguet-Boreu, Gemma Flores-Mateo, Chris Salisbury, Jeanet Blom, Michael Freitag, Liam Glynn, Christiane Muth, and Jose M. Valderas. Prevalence, determinants and patterns of multimorbidity in primary care: A systematic review of observational studies. PLOS One, 9(7), JUL 21 2014.

[6] S. N. Etkind, A. E. Bone, B. Gomes, N. Lovell, C. J. Evans, I. J. Higginson, and F. E. M. Murtagh. How many people will need palliative care in 2040? past trends, future projections and implications for services. BMC Medicine, 15, MAY 18 2017.

[7] F. Alhasoun, F. Aleissa, M. Alhazzani, L. G. Moyano, C. Pinhanez, and M. C. Gonzalez. Age density patterns in patients medical conditions: A clustering approach. PLOS Computational Biology, 14(6), 2018. Alhasoun, Fahad Aleissa, Faisal Alhazzani, May Moyano, Luis G. Pinhanez, Claudio Gonzalez, Marta C. Gonzalez, Marta C./E-9456-2011 Gonzalez, Marta C./0000-0002-8482-0318 1553-7358.

[8] I. Ioakeim-Skoufa, B. Poblador-Plou, J. Carmona-Pirez, J. Diez-Manglano, R. Navickas, L. A. Gimeno-Feliu, F. Gonzalez-Rubio, E. Jureviciene, L. Dambrauskas, A. Prados-Torres, and A. Gimeno-Miguel. Multimorbidity patterns in the general population: Results from the epichron cohort study. International Journal of Environmental Research and Public Health, 17(12), 2020.

[9] Miika Koskinen, Jani K. Salmi, Anu Loukola, Mika J. Makela, Juha Sinisalo, Olli Carpen, and Risto Renkonen. Data-driven comorbidity analysis of 100 common disorders reveals patient subgroups with differing mortality risks and laboratory correlates. Scientific Reports, 12(1), NOV 2 2022.

[10] Richard M. Dodds, Jonathan G. Bunn, Susan J. Hillman, Antoneta Granic, James Murray, Miles D. Witham, Sian M. Robinson, Rachel Cooper, and Avan A. Sayer. Simple approaches to characterising multiple long-term conditions (multimorbidity) and rates of emergency hospital admission: Findings from 495,465 uk biobank participants. Journal of Internal Medicine, 2022.

[11] Ignatios Ioakeim-Skoufa, Mercedes Clerencia-Sierra, Aida Moreno-Juste, Carmen Elias de Molins Pena, Beatriz Poblador-Plou, Mercedes Aza-Pascual-Salcedo, Francisca Gonzalez-Rubio, Alexandra Prados-Torres, and Antonio Gimeno-Miguel. Multimorbidity clusters in the oldest old: Results from the epichron cohort. International Journal of Environmental Research and Public Health, 19(16), AUG 2022.

[12] A. J. Webster and R. Clarke. Sporadic, late-onset, and multistage diseases. PNAS Nexus, 1(3), 2022.

[13] Clare Bycroft, Colin Freeman, Desislava Petkova, Gavin Band, Lloyd T. Elliott, Kevin Sharp, Allan Motyer, Damjan Vukcevic, Olivier Delaneau, Jared O Connell, Adrian Cortes, Samantha Welsh, Alan Young, Mark Effingham, Gil McVean, Stephen Leslie, Naomi Allen, Peter Donnelly, and Jonathan Marchini. The UK Biobank resource with deep phenotyping and genomic data. Nature, 562(7726):203–209, 2018.

[14] P. Armitage and R. Doll. The age distribution of cancer and a multi-stage theory of carcinogenesis. British Journal of Cancer, 8(1):1–12, 1954.

[15] P. Armitage. Multistage models of carcinogenesis. Environmental Health Perspectives, 63(NOV):195–201, 1985.

[16] Anthony J. Webster. Multi-stage models for the failure of complex systems, cascading disasters, and the onset of disease. PLOS One, 14(5), 2019.

[17] Y.H. Wang. On the number of successes in independent trials. Statistica Sinica, 3:295–312, 1993.

[18] Yili Hong. On computing the distribution function for the poisson binomial distribution. Computational Statistics and Data Analysis, 59:41–51, 2013.

[19] A. J. Webster. Causal attribution fractions, and the attribution of smoking and bmi to the landscape of disease incidence in uk biobank. Scientific Reports, 12(19678), 2022.

[20] Rounak Dey, Ellen M. Schmidt, Goncalo R. Abecasis, and Seunggeun Lee. A fast and accurate algorithm to test for binary phenotypes and its application to phewas. The American Journal of Human Genetics, 101(1):37–49, 2017.

[21] L. Wasserman. All of Statistics. Springer, 2005.

[22] World Health Organization. International statistical classification of diseases and related health problems 10th revision, 2016.

[23] A. J. Webster, K. Gaitskell, I. Turnbull, B. J. Cairns, and R. Clarke. Characterisation, identification, clustering, and classification of disease. Scientific Reports, 11(1), 2021.

[24] Webster, A. J. Gaitskell, K. Turnbull, I. Cairns, B. J. Clarke, R. J. Pearl. Causality. John Wilely & Sons Ltd, 2nd edition edition, 2009.

[25] J. Pearl, M. Glymour, and N.P. Jewell. Causal Inference In Statistics. Cambridge University Press, 1st edition edition, 2016.

[26] T.J. VanderWeele. Explanation in Causal Inference. Oxford University Press, 1st edition edition, 2015.

[27] Terry M Therneau. A Package for Survival Analysis in R, 2023. R package version 3.5-0.

[28] Craig Varrichio. grr: Alternative Implementations of Base R Functions, 2016. R package version 0.9.5.

[29] Matt Dowle and Arun Srinivasan. data.table: Extension of ‘data.frame’, 2021. R package version 1.14.0.

[30] Arne Henningsen and Ott Toomet. maxlik: A package for maximum likelihood estimation in R. Computational Statistics, 26(3):443–458, 2011.

